# COVID-19 vaccination hesitancy model: The impact of vaccine education on controlling the outbreak in the United States

**DOI:** 10.1101/2021.05.21.21257612

**Authors:** Bismark Oduro, Attou Miloua, Ofosuhene O. Apenteng, Prince P. Osei

**Author notes:** Corresponding author: Department of Mathematics and Physical Sciences, California University of PA, California, PA 15419, USA.

## Abstract

The coronavirus outbreak continues to pose a significant challenge to human lives globally. Many efforts have been made to develop vaccines to combat this virus. However, with the arrival of the COVID-19 vaccine, there is hesitancy and a mixed reaction toward getting the vaccine. We develop a mathematical model to analyze and investigate the impacts of education on individuals hesitant to get vaccinated. The findings indicate that vaccine education can substantially minimize the daily cumulative cases and deaths of COVID-19 in the United States. The results also show that vaccine education significantly increases the number of willing susceptible individuals, and with a high vaccination rate and vaccine effectiveness, the outbreak can be controlled in the US.

## 1 Introduction

The severe acute respiratory syndrome coronavirus (SARS–CoV–2) strain that caused 2019 novel coronavirus disease (COVID-19) pandemic was first observed in Wuhan, China, in December 2019 and was later declared a pandemic by the World Health Organization (WHO) on March 11, 2020 [1]. The first case of coronavirus in the United States of America was revealed on January 20, 2020, in Washington State. Since then, the number of positive cases has reached more than 30 million across the country in just over one year. COVID-19 is spread from person to person mainly through respiratory droplets released when an infected person sneezes, coughs or speaks [1, 2]. Several non-pharmaceutical measures, including face mask use, social distance, quarantining, etc., are recommended to reduce the virus’s spread. However, there is still a need for public health and clinical interventions to successfully contain the disease.

Health experts agree that the best way to end the pandemic is to vaccinate most of the population [3, 4]. Currently, three COVID-19 vaccines are already known, recommended, and being used in the USA. The first is the Pfizer-BioNTech; one must complete two doses, about three weeks apart, recommended for individuals aged 16 years and older. The second vaccine available is called Moderna COVID-19 vaccine. It is also to be taken in two doses, one month or 28 days apart, introduced by a shot in the muscle of the upper arm. It is recommended for people aged 18 years and older. The third vaccine is called Johnson and Jonson’s Janssen COVID-19 vaccine, got emergency use authorized on February 27, 2021. These vaccines are intended for the prevention of coronavirus disease. They may help to mitigate the spread of the coronavirus once most individuals make an effort to be vaccinated. However, a large proportion of the American population is reluctant about the COVID-19 vaccines. Lack of information about the side effects, especially the long-term effects, time-line of the COVID vaccines production, political [5] and conspiracy theory, are among the reasons for vaccine hesitancy.

The population opinion and the trust in the vaccine are of the most significant importance for appropriate coverage. A report in the American Medical Association Journal shows that skepticism toward the COVID-19 Vaccines is on the rise among Americans. A survey by the Kaiser Family Foundation shows that about 29% of health workers were hesitant to accept the vaccine [6]. In [7], a sample of 1878 adult Americans was asked different questions related to vaccination. The sample is composed of (52%) Females, (74%) Whites, (81%) non-Hispanic, (56%) married, (68%) employed full time, (77%) with a bachelor’s degree and higher. The probability of receiving the vaccine in the research was as follows: (52%) for very likely, (27%) for somewhat likely, (15%) not likely, (7%) definitely not. Vaccine hesitancy was also higher in African Americans, Hispanics, and pregnant women, and breastfeeding moms.

Mathematical models are powerful tools for investigating human infectious diseases, such as COVID-19, contributing to the understanding of infections’ dynamics, and can provide valuable information for public-health policymakers [8, 9]. Numerous mathematical models have been used to provide insights into public health measures for mitigating the spread of the coronavirus pandemic [10, 11, 12, 13]. For example, Eikenberry et al. in [14] developed a mathematical model to assess the impact of mask use by the general public on the transmission dynamics of the COVID-19 pandemic. Their results showed that broad adoption of even relatively ineffective face masks might reduce community transmission of COVID-19 and decrease peak hospitalizations and deaths.

Given the pervasiveness of vaccine hesitancy, we develop a compartmental model to study the impacts of education for individuals unwilling to accept the COVID-19 vaccines. We explore policy-related questions, including investigating the vaccination rate and vaccine education impact on disease dynamics in the USA.

## 2 Model formulation

We consider a compartmental model for the infection’s transmission dynamics and control. With the arrival of various COVID–19 vaccines, there is a mixed reaction to get vaccinated or not. We classify the US’s total population into two subgroups: Those willing to get the vaccine and those unwilling to receive the vaccine. We further sub-divide the populations into eleven mutually exclusive compartments of willing susceptible (*S*_*w*_), unwilling susceptible (*S*_*u*_), willing early-exposed (*E*_*ew*_), unwilling early-exposed (*E*_*eu*_), willing late-exposed (*E*_*lw*_), unwilling late-exposed (*E*_*lu*_), willing infected (*I*_*w*_), unwilling infected (*I*_*u*_), willing recovered (*R*_*w*_), unwilling recovered (*R*_*u*_), and vaccinated population (*V*) so that the total population at time *t*, denoted by*N* (*t*) is

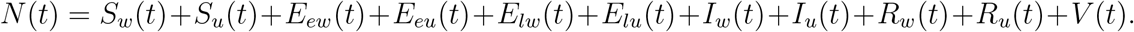

We incorporate education for the unwilling populations at a rate *σ*. The willing populations receive the vaccine at rate *r* and progress to the vaccinated class. There is evidence that individuals exposed or infected should wait for 90 days before receiving the COVID-19 vaccine; we exclude exposed and infected individuals from the vaccination until they are recovered. Figure 1 fully illustrates the flow of populations in the various compartments; the model’s parameters and their descriptions are listed in Table 2.

**Table 1:** Description of state variables of the COVID-19 model.

| State variable | Description |
| --- | --- |
| $S_w(S_u)$ | The population of willing (unwilling) susceptible individuals |
| $E_{ew}(E_{eu})$ | The population of willing (unwilling) early-exposed individuals (i.e, newly-infected individuals who are not yet infectious) |
| $E_{lw}(E_{lu})$ | The population of willing (unwilling) late-exposed individuals (i.e., exposed individuals who are close to surviving the incubation period and are shedding virus) |
| $I_w(I_u)$ | The population of willing (unwilling) infected individuals |
| $R_w(R_u)$ | The population of willing (unwilling) recovered individuals |
| $V$ | The population of vaccinated individuals |

**Table 2:**
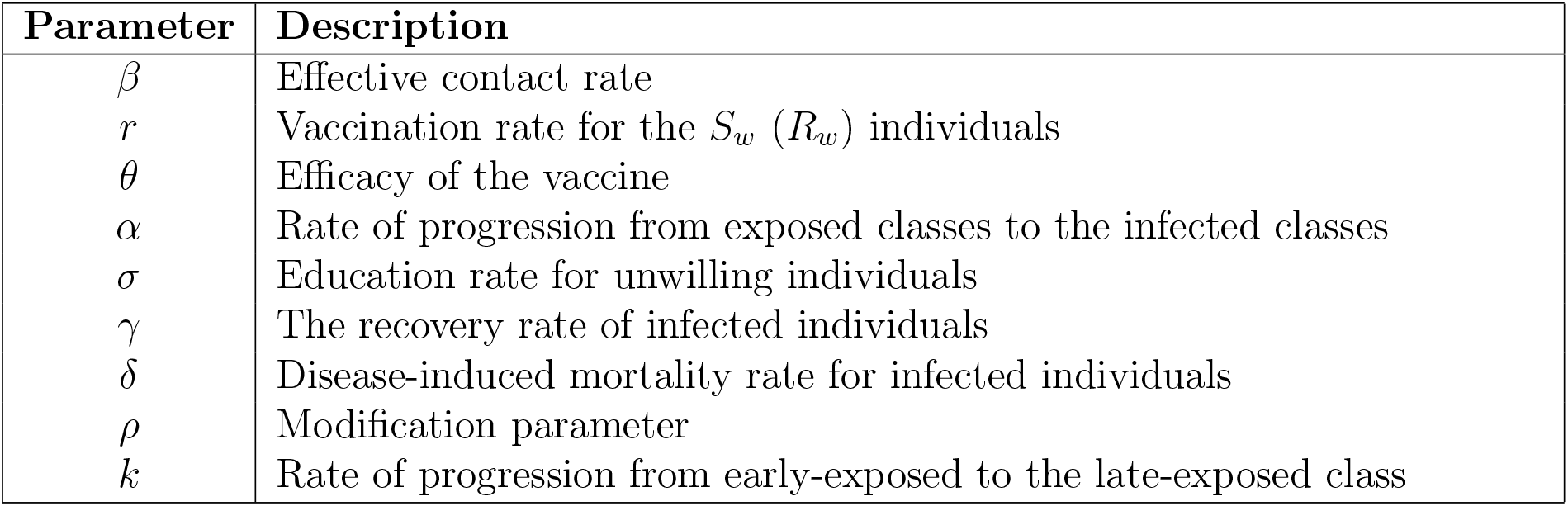
Description of the parameters of the COVID-19 model (1).

| Parameter | Description |
| --- | --- |
| $\beta$ | Effective contact rate |
| $r$ | Vaccination rate for the $S_w$ ($R_w$) individuals |
| $\theta$ | Efficacy of the vaccine |
| $\alpha$ | Rate of progression from exposed classes to the infected classes |
| $\sigma$ | Education rate for unwilling individuals |
| $\gamma$ | The recovery rate of infected individuals |
| $\delta$ | Disease-induced mortality rate for infected individuals |
| $\rho$ | Modification parameter |
| $k$ | Rate of progression from early-exposed to the late-exposed class |

**Figure 1:**
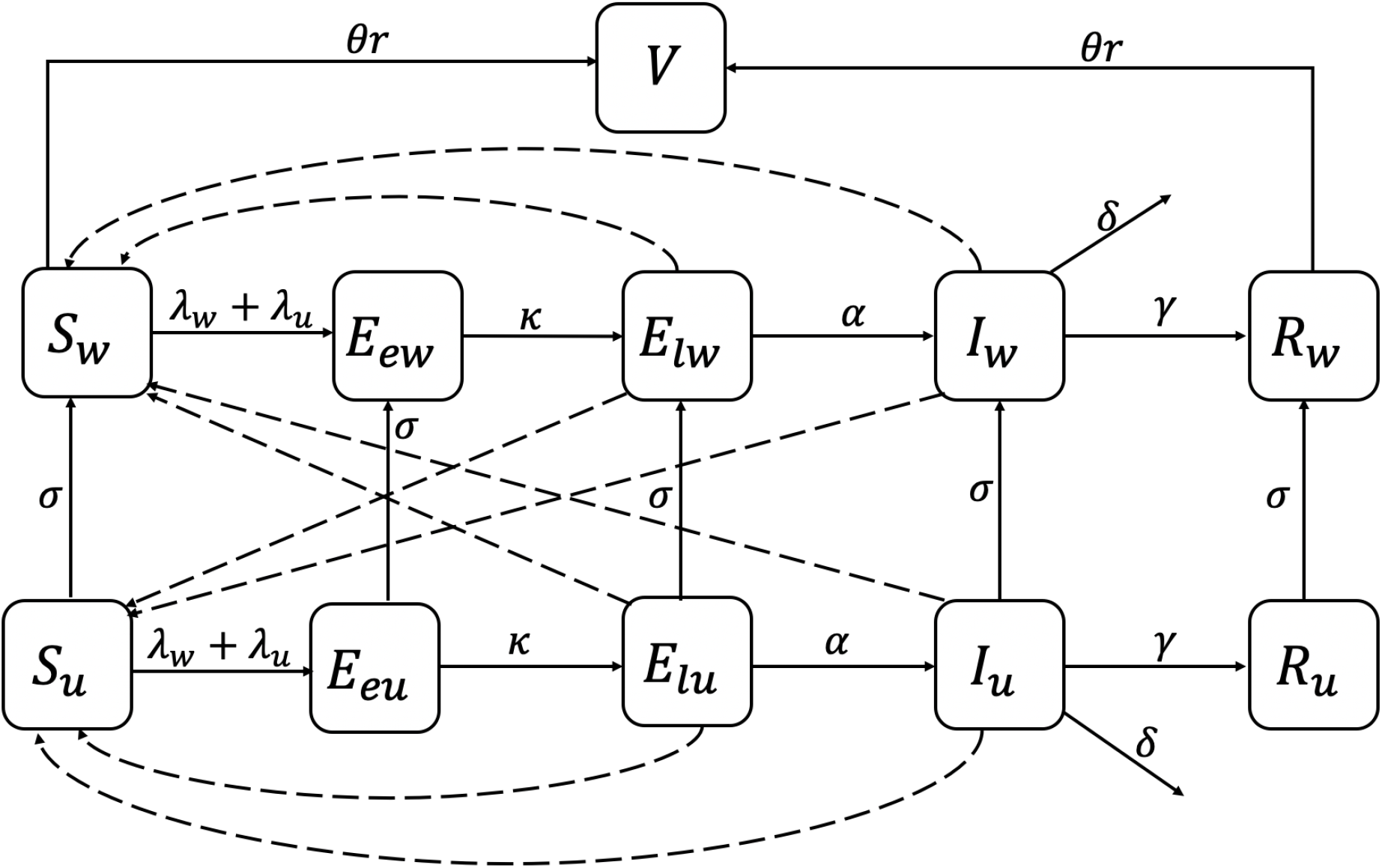
Schematic diagram of the COVID–19 model

The dynamics in Figure 1 can be represented as a system of nonlinear ordinary differential equations given by

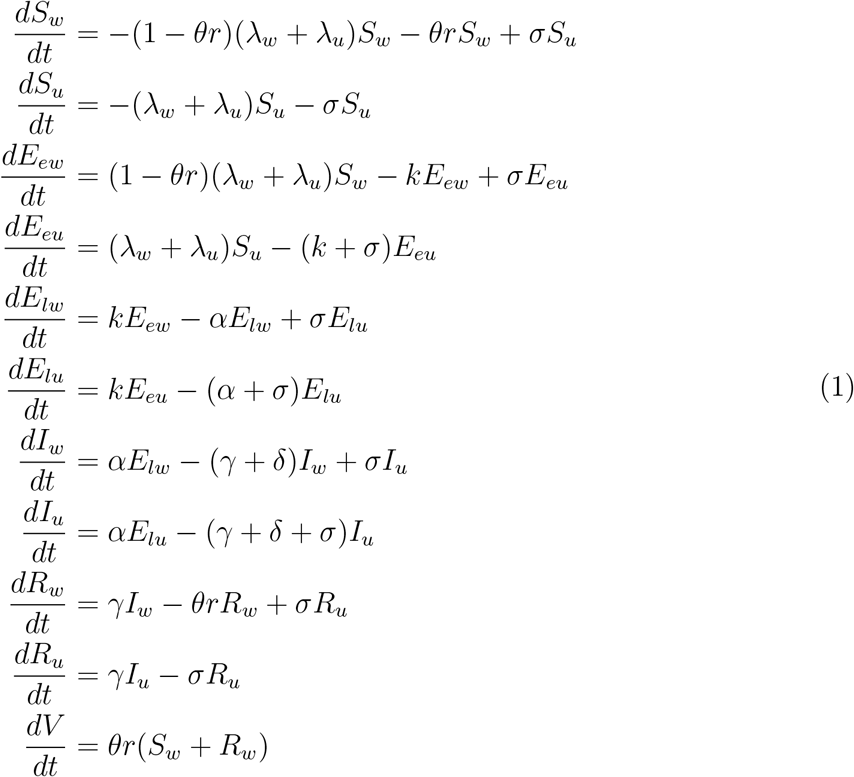

where the forces of infection are given by

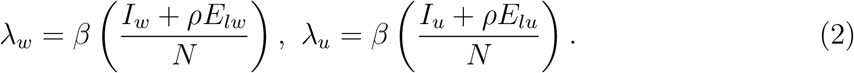

## 3 Results

### 3.1 Disease-free equilibrium and reproduction number

The model (1) has a disease-free equilibrium (DFE) given by

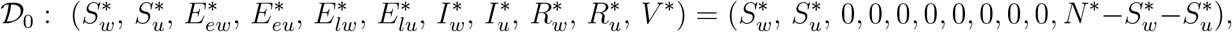

where *N* ^*∗*^ is the initial total population size, 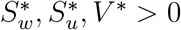, and 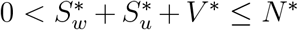. The next generation operator method [15, 16, 17] can be used to analyze the asymptotic stability property of the disease-free equilibrium, 𝒟_0_. In particular, using the notation in [15, 16, 17], it follows that the associated next generation matrices, *F* and *V*, for the new infection terms and the transition terms, are given, respectively, by

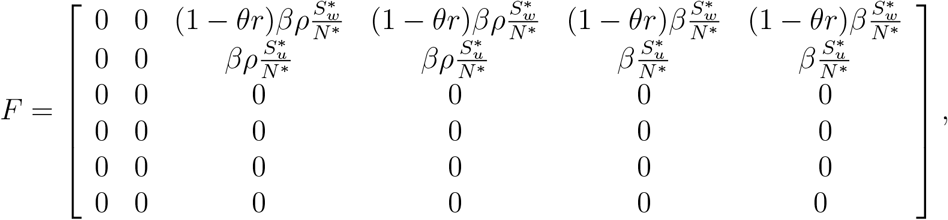

and,

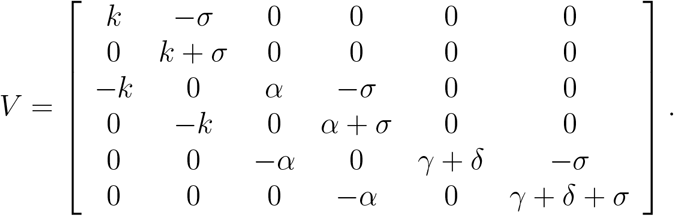

The control reproduction number is given by

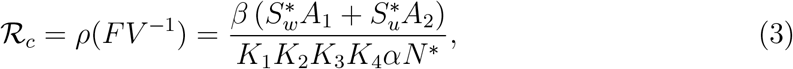

Where

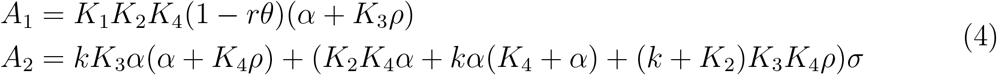

with *K*_1_ = *k* + *σ, K*_2_ = *α* + *σ, K*_3_ = *γ* + *δ*, and *K*_4_ = *γ* + *δ* + *σ*.

The reproduction number is the average number of new COVID-19 cases generated by a typical infectious individual introduced into a population where a certain fraction is protected; it is a measure of contagiousness of infectious diseases.

#### Theorem 3.1

*The disease-free equilibrium (DFE) of the model* (1) *is locally-asymptotically stable if ℛ*_*c*_ *<* 1. *When ℛ*_*c*_ *>* 1, *we will initially observe a near exponential growth of infectious cases, reach a peak, and eventually declines to an equilibrium*.

### 3.2 Parameter estimation

The proposed model is fitted and validated using the USA COVID–19 daily cumulative cases and deaths from December 14, 2020, to January 20, 2021. The choice of this data is motivated by the commencement of national vaccination on December 14, 2020, in the USA. There are nine parameters underlying the model; however, three of the parameters, i.e., *γ, δ*, and *θ*, were fixed, and the rest were estimated. It is worth noting that the model parameters may not be uniquely identifiable based only on cumulative cases and deaths of USA COVID–19 data available. We addressed this identifiability problem by using an inverse modelling, sensitivity and Monte Carlo analysis method built in FME [18] package in R (see also [19]). This method analyzes mathematical models with data, performs local and global sensitivity, and Monte Carlo analysis. It addresses parameter identifiability issues and fits a model to data using existing optimization methods such as the constrained quasi-Newton method.

The estimated parameter values obtained from the best model prediction are given in Table 3. The fitted models and the observed data for the coronavirus cumulative cases and deaths in the USA are displayed in Figure 2. We observe that the model fit the observed data significantly.

**Table 3:** Estimated parameter values for the model (1) using data for the US.

| Parameters | 14/12/2020–20/1/2021 | Source |
| --- | --- | --- |
| $\beta$ | 0.1519 | Estimated |
| $r$ | 0.0131 | Estimated |
| $\alpha$ | 0.5000 | Estimated |
| $\sigma$ | 0.6397 | Estimated |
| $\rho$ | 0.2230 | Estimated |
| $k$ | 0.7550 | Estimated |
| $\gamma$ | 0.1 | [20] |
| $\delta$ | 0.0015 | [20] |
| $\theta$ | 0.9469 | Assumed |

**Figure 2:**
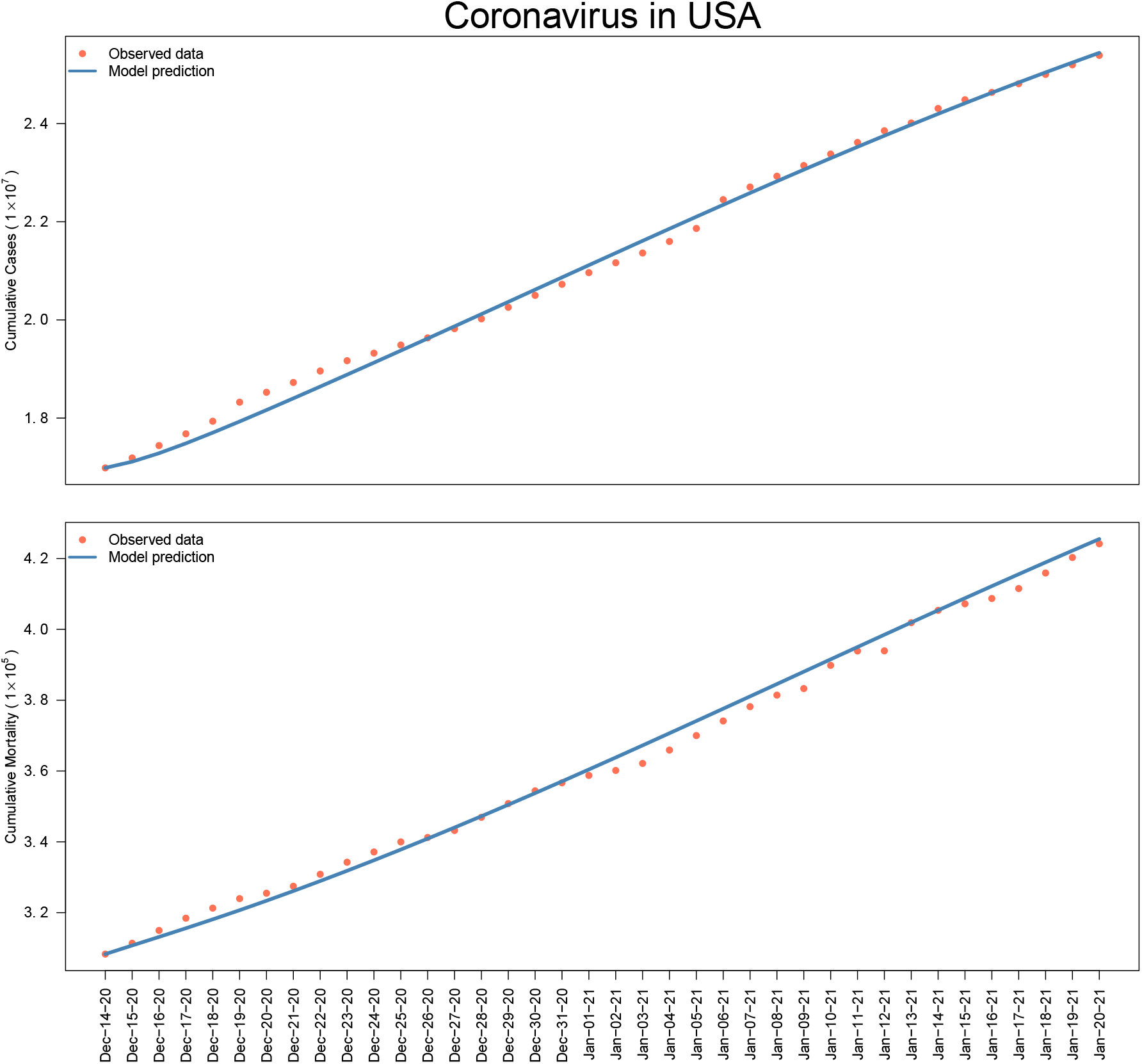
The fitted model and the observed COVID–19 cumulative cases and deaths in USA from December 14, 2020 to January 20, 2021

### 3.3 Sensitivity analysis

This section uses the Latin Hypercube Sampling and Partial Rank Correlation Coefficients (PRCC) to perform sensitivity analysis [21, 22] on the model parameters. The analysis is needed to identify model parameters that significantly impact model outcomes using the reproduction number (ℛ_*c*_) as the response function [12, 21, 22]; that is, to determine the model robustness to parameter values. Parameters with large PRCC greater than +0.50 are strongly positively correlated with the response function. In contrast, those less than *−*0.50 are said to be largely negatively associated with the response function [21, 22]. Figure 3 displays the PRCC analysis plot of the model parameters considered. The results show that the effective contact rate (*β*) and the modification parameter (*ρ*) positively affect ℛ_*c*_, meaning that an increase in these parameters will increase ℛ_*c*_. On the other hand, the education rates for unwilling individuals (*σ*), the vaccination rate *r*, and vaccine efficacy (*θ*) have a negative effect on the *ℛ*_*c*_, and increasing these parameters would lower the *ℛ*_*c*_. The results further indicate that the parameters; the effective contact rate, education rate for unwilling individuals, the vaccination rate, and the vaccine efficacy mainly influence the response function (*ℛ*_*c*_).

**Figure 3:**
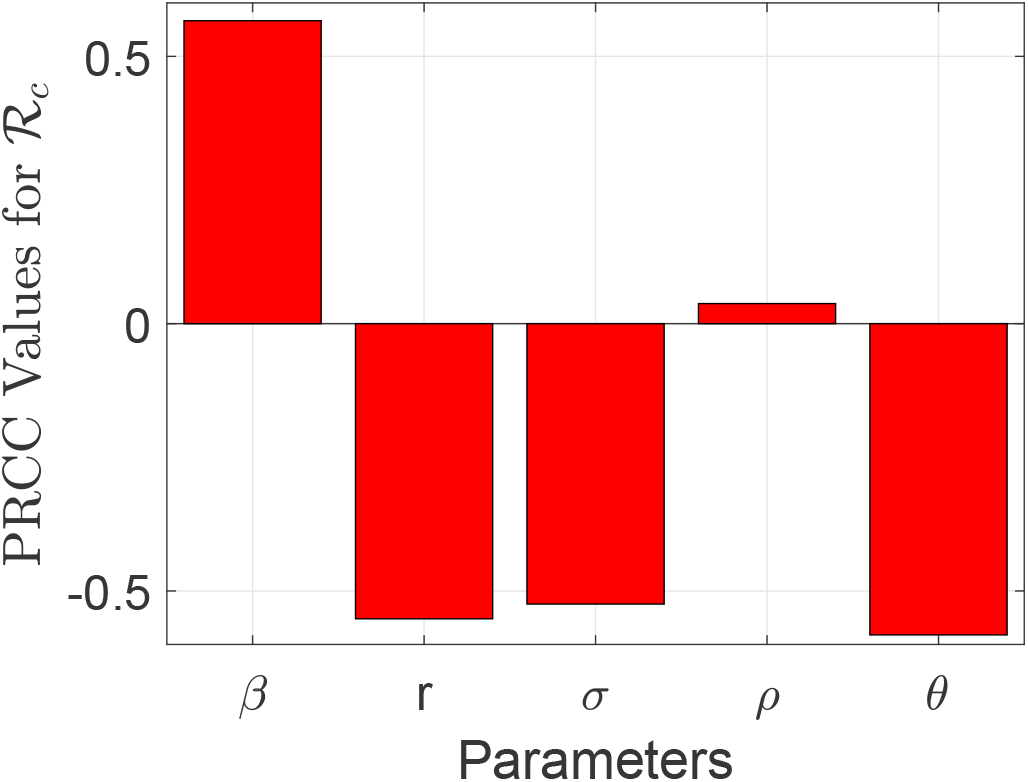
Partial rank correlation coefficients (PRCCs) showing the impact of model parameters on the reproduction number (ℛ_*c*_) of the model. Parameter values used are as given in Table 3.

### 3.4 Contour plots results

We generated contour plots to analyze the reproduction number (ℛ_*c*_) of the model as a function of desired parameters as displayed Figure 4. Parameter values used for the simulation are as given in Table 3.

**Figure 4:**
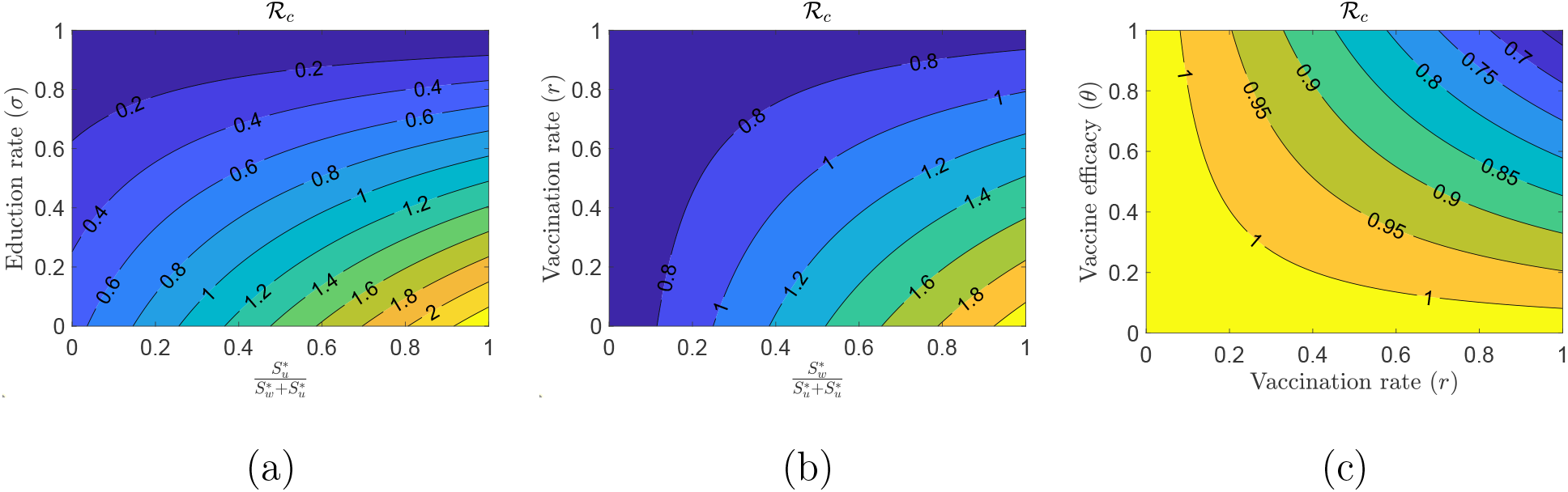
Contour plot of the reproduction number (*ℛ*_*c*_) of the model (1), as a function of: (a) Education rate (*σ*) and the ratio 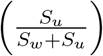, (b) Vaccination rate (*r*) and the ratio 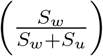, and (c) Vaccination efficacy (*θ*) and vaccination rate (*r*).

Based on the contour plot results in Figure 4, the following observation inferences are made:

i. To effectively curb the outbreak, that is, reducing (ℛ _*c*_) to a value less than unity, the Figure 4 (a) suggests that a high education rate (*σ*) is needed. The lower the fraction of individuals hesitant to accept the vaccine, the lower the education rate required and vice versa. For example, to decrease the reproduction number below one, at least an education rate of 0.3 is needed if about 40% of the susceptible population is unwilling to vaccinate.
ii. The ℛ_*c*_ decreases as more individuals are being educated and are willing to receive the vaccine, and the vaccination rate is high, Figure 4 (b).
iii. Figure 4 (c) illustrates the impact of a relatively high vaccination rate and efficacy. One can observe that a high combined effects cause the *ℛ* _*c*_ to drop to below one.

### 3.5 Time evolution analysis, and predictions

In this section, we analyze the effects of vaccine education *σ*, the vaccination rate *r*, and the efficacy of the vaccine *θ*, on the cumulative US COVID-19 cases and deaths. In addition, we provide some future predictions of the cumulative cases and deaths with their associated uncertainty.

First, we begin with the effects of the education for unwilling individuals and its impact on the cumulative cases and deaths. Table 4 provides numerical description while Figure 5 graphically explains the decrease and increase in cumulative cases and deaths when the education rate is increased or decreased, respectively. For example, from Table 4, a 25% increase in education rate reduces the cumulative cases from 25441752 to 25108128 and from 425513 to 422423 for the cumulative deaths.

**Table 4:** A summary of various increase in education rate.

| **Education rate ($\sigma$)** | **14/12/2020–20/1/2021** |  |
| --- | --- | --- |
|  | Cumulative Cases | Cumulative Deaths |
| Baseline | 25441752 | 425513 |
| 25% increase | 25108128 | 422423 |
| 25% decrease | 25938095 | 430248 |
| 50% increase | 24832044 | 419893 |
| 50% decrease | 27393963 | 444084 |

**Figure 5:**
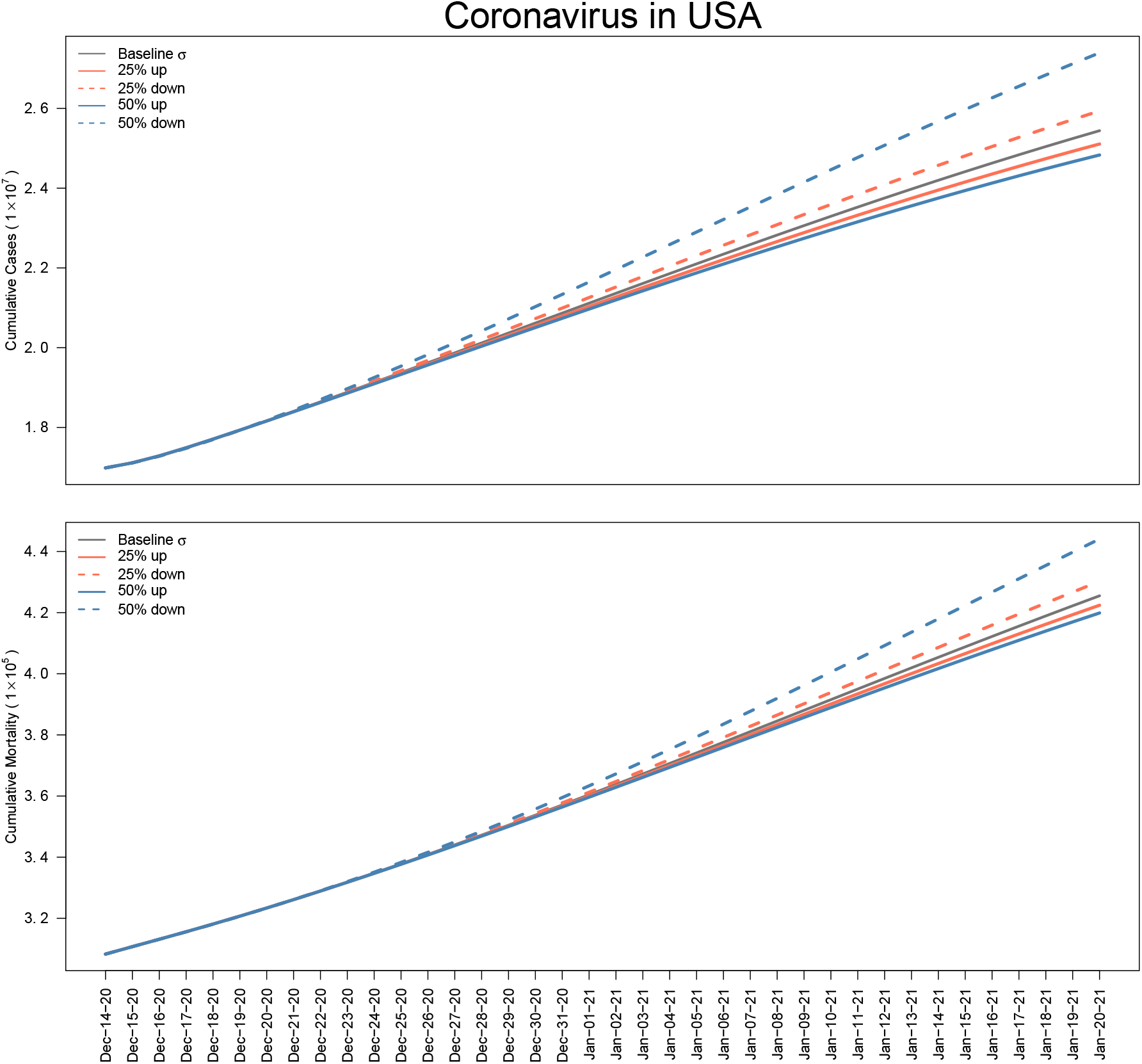
Effects of increasing and decreasing the baseline education rate parameter *σ*

We observe that an increase in vaccine education decreases the COVID-19 cumulative cases and deaths. On the other hand, a lower vaccine education increases the baseline cumulative cases and deaths. This suggest that, an increase of education parameter from the baseline, lowers the spread of COVID-19. It is worth noting that while the curves for the baseline education parameter and its increase or decrease overlap up until the end of the year 2020, they are separated beginning of January 2021.

Second, we analyze the effects of increasing or decreasing the vaccination rate *r*, on cumulative cases and deaths numerically and graphically. The results are displayed in Table 5 and Figure 6.

**Table 5:** A summary of various increase in vaccination rate.

| Vaccination rate ($r$) | 14/12/2020–20/1/2021 | |
| --- | --- | --- |
|  | Cumulative Cases | Cumulative Deaths |
| Baseline | 25441752 | 425513 |
| 25% increase | 24771267 | 419830 |
| 25% decrease | 26223401 | 431946 |
| 50% increase | 24192245 | 414785 |
| 50% decrease | 27141116 | 439270 |

**Figure 6:**
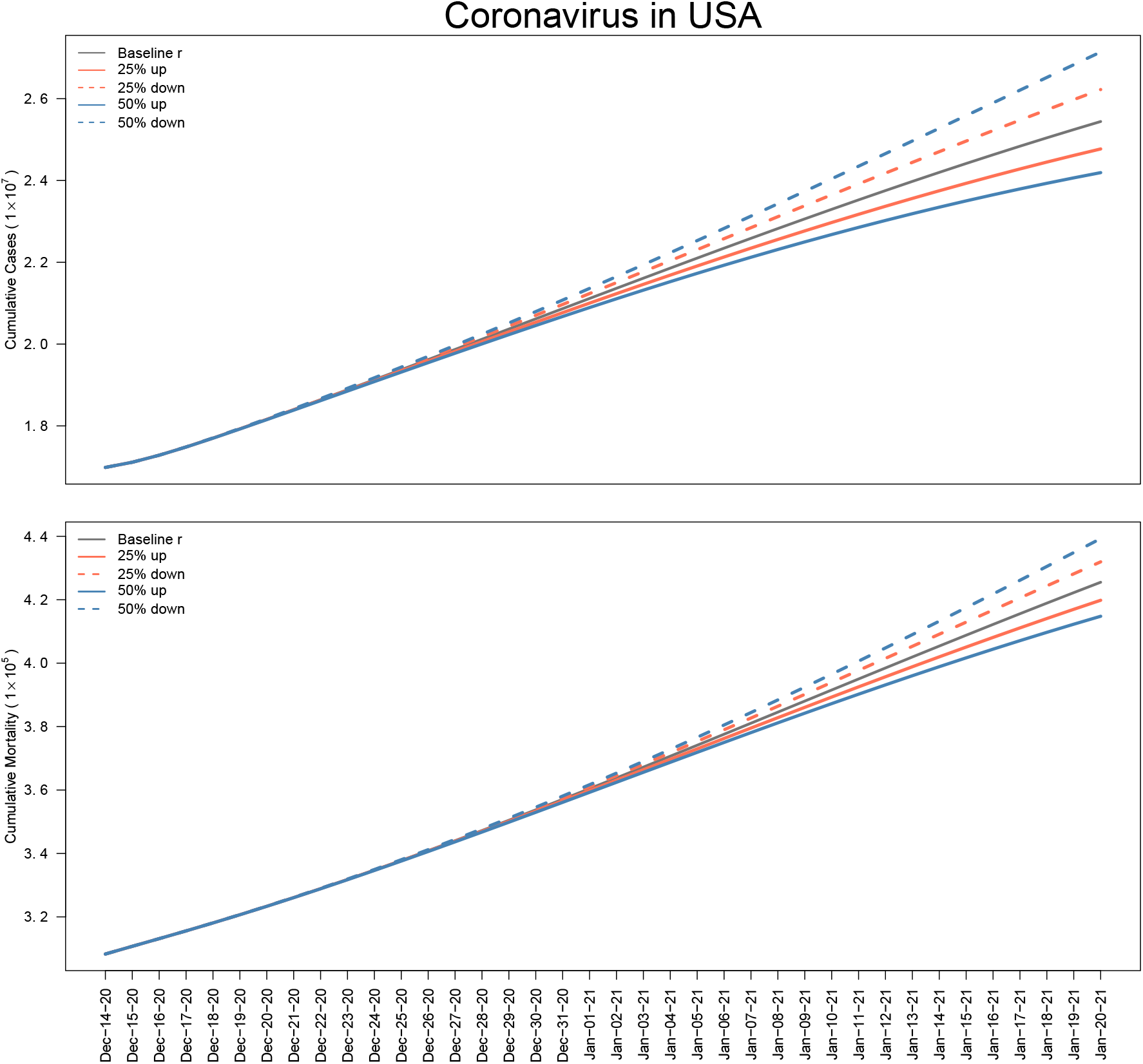
Effects of increasing and decreasing the baseline of vaccination rate, *r*

Note that 50% increase or decrease in the vaccination rate decreases or increases the cumulative cases and deaths significantly compared to the baseline scenario. For instance, a 50% increase in the baseline rate of vaccination reduces the cumulative cases and deaths from 25441752 and 425513 to 24192245 and 414785 respectively. On the other hand, the same reduction rate from the baseline increases the cumulative cases and deaths to 27141116 and 439270, respectively. Note also that the 50% increase of the vaccination rate is significantly below the baseline after January 15, 2021, while a 50% reduction of the rate of vaccination is significantly above the baseline curve. This suggests that as the vaccination rate increases, the cumulative cases and deaths of COVID–19 in the USA decreases.

Finally, we consider the effects of the efficacy of the vaccine and report the numerical and graphical results of varying the parameter *θ*. Note that the baseline vaccine efficacy was high, and thus the maximum increment adopted in this communication stands at 5%, which represents 99.4% efficacy. The quantitative results and graphical description of various decrease and an increase in the efficacy of the vaccine is displayed in Table 6 and Figure 7.

**Table 6:** A summary of various decrease and an increase in the efficacy of COVID–19 vaccine.

| **Efficacy of the vaccine ($\theta$)** | **14/12/2020–20/1/2021** |  |
| --- | --- | --- |
|  | Cumulative Cases | Cumulative Deaths |
| Baseline | 25441752 | 425513 |
| 5% increase | 25299458 | 424320 |
| 45% decrease | 26945280 | 437726 |
| 35% decrease | 26572557 | 434760 |
| 25% decrease | 26223401 | 431946 |

**Figure 7:**
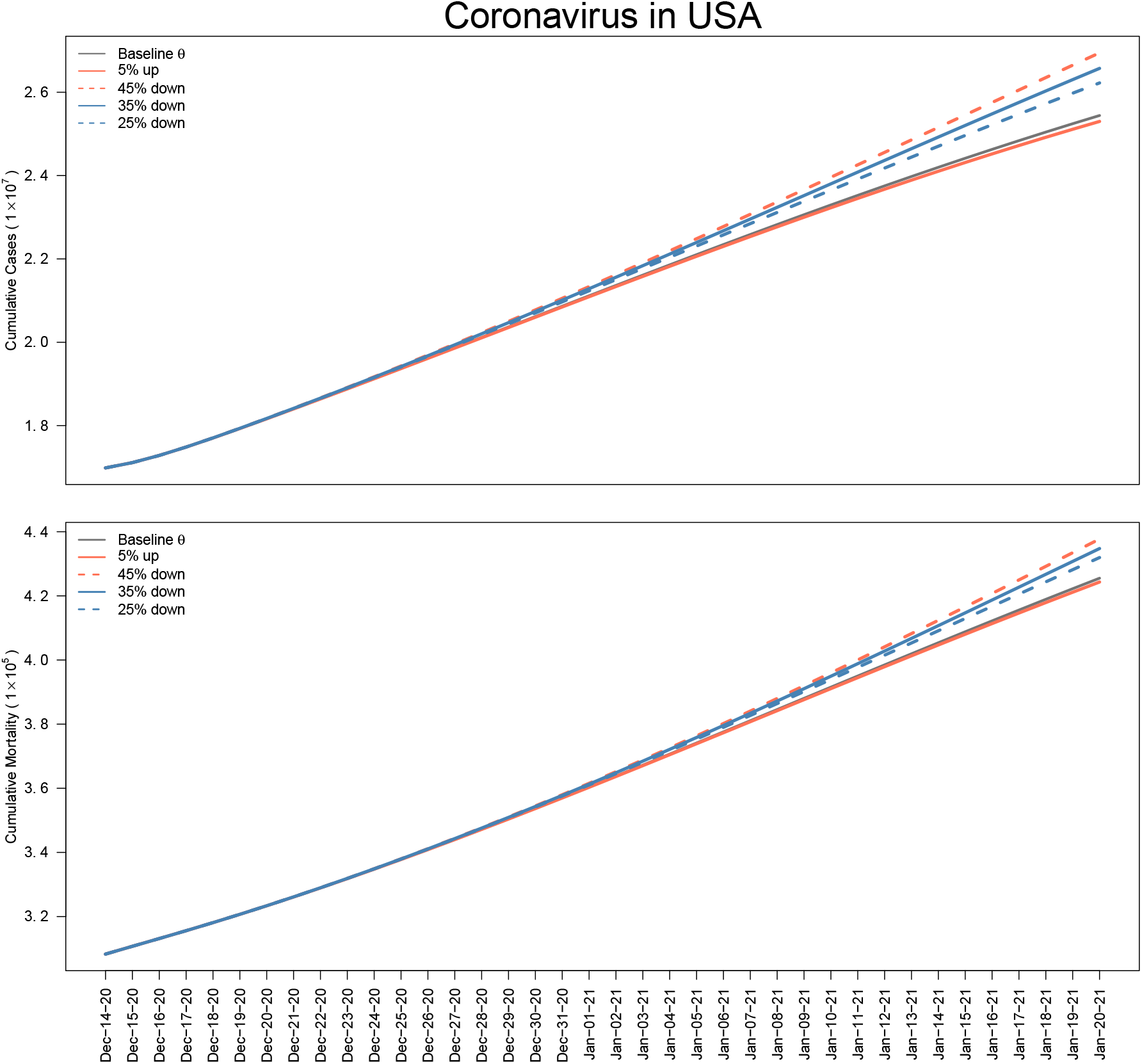
Effects of increasing and decreasing the baseline efficacy of the COVID–19 vaccine *θ*

We observe that a lower vaccine efficacy has the highest cumulative cases and deaths of COVID-19. Note that a 45% reduction of vaccine efficacy from the baseline, i.e., a 52.1% vaccine efficacy increases the cumulative cases and deaths from the baseline 25441752 to 26945280 and 425513 to 437726, respectively. Also, note that the curve of the 5% increase of the baseline efficacy of the vaccine is similar to the baseline line.

We validate our model by predicting the cumulative cases and deaths every two weeks after January 20, 2021 for the USA COVID–19 outbreak; all the data are given in the format of 95% Confidence Interval (CI) of the predicted values. The uncertainty of the predicted values are characterized using the the 95% confidence intervals (CI). We compared the estimated predicted values to the actual values reported in [23] within the 95% CI, as shown in Table 7. Our model is robust and can be replicated for other countries.

**Table 7:** Prediction and reported values.

| Cumulative | Date | Predicted | 95% CI | Reported[23] |
| --- | --- | --- | --- | --- |
| Cases | Feb. 3, 2021 | 28218225 | (24890729 - 31545721) | 27216525 |
|  | Feb. 17, 2021 | 30994706 | (21977405 - 40012007) | 28544574 |
|  | Mar. 3, 2021 | 33771187 | (17445706 - 50096669) | 29502980 |
|  | Mar. 17, 2021 | 36547669 | (11597638 - 61497699) | 30299662 |
|  | Mar. 31, 2021 | 39324150 | (4610138 - 74038162) | 31166344 |
| Deaths | Feb. 3, 2021 | 471357 | (465599 - 477116) | 469431 |
|  | Feb. 17, 2021 | 517202 | (500904 - 533500) | 504302 |
|  | Mar. 3, 2021 | 563047 | (532582 - 593512) | 532807 |
|  | Mar. 17, 2021 | 608891 | (560978 - 656805) | 551583 |
|  | Mar. 31, 2021 | 654736 | (586236 - 723236) | 565256 |

## 4 Discussion and conclusions

The coronavirus outbreak poses a severe threat to human lives globally. Since the beginning of the pandemic, a campaign to use non-pharmaceutical measures to prevent the virus’s spread has been underway, including a face mask, social distancing, and frequent hand washing. The emergence of vaccines sounds promising; however, there is a challenge of vaccine hesitancy. It is therefore essential to understand how educating the general public will impact the fight against the outbreak. In this paper, we use a compartmental model with vaccine education to study the dynamics of the COVID-19 infection. We classify the US’s total population into two subgroups: Those willing to accept the vaccine and those unwilling to receive the vaccine. The vaccine education is incorporated for the general public hesitant to take the vaccine. We assessed the impact of the education campaign on the control of the outbreak.

First, we computed an expression for the reproduction number (ℛ_*c*_), a threshold that measures the contagiousness of infectious diseases. We performed a sensitivity analysis of the reproduction number. The result shows that vaccine education negatively influences the reproduction number; that is, an increase in the vaccine education implies a decrease in the ℛ_*c*_. Epidemiologically, when ℛ_*c*_ *≤*1, then the transmission will fade or die out. In contrast, the infected number of people is expected to increase if ℛ_*c*_ *>* 1. The sensitivity analysis result also shows that vaccine efficacy and vaccination rate negatively impact theℛ_*c*_, and raising them will reduce the ℛ_*c*_.

Using contour plots, we further analyzed the reproduction number as a function of two independent variables, the vaccine education and the proportion of unwilling susceptible individuals 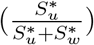. The prospect of curtailing the outbreak is achievable with a high education rate. For instance, the result in Figure 4 (a) shows that if a high proportion of the susceptible populations is unwilling to accept the vaccine, high public education can help diminish the reproduction number and mitigate the virus. Also, vaccine education raises the willing susceptible individuals, and with a high vaccination rate and vaccine efficacy, it will be possible to control the spread of the outbreak.

Next, we analyze the effects of vaccine education, vaccination rate, and the efficacy of the vaccine on the daily cumulative cases and deaths from December 14, 2020 to January 20, 2021. The results suggest that higher rates of vaccine education, vaccination, and vaccine efficacy contribute to the mitigation of the spread of the COVID-19 disease outbreak. For example, the result in Figure 5 shows that a lower vaccine education contributes to the higher projected cumulative cases and deaths. On the other hand, higher education rates help to control the outbreak. The effect of these higher rates becomes evident over time. These results suggest that a combined effect of higher vaccine education, vaccination, and vaccine efficacy rates would contribute to slow the pace of the spread of the COVID-19 outbreak and thus help the government to control the disease. The mitigation effect of higher rates of education, vaccination, and vaccine efficacy is clearly evident over time as we observe from the curves in Figures 5, 6 and 7; thus, these mitigation strategies need to be enforced consistently over time.

Finally, to validate and test the robustness of our proposed mathematical model, we predicted the cumulative cases and deaths every two weeks after January 20, 2021. The reported cumulative cases and fatalities in Table 7 almost all fall within the prediction interval of 95% confidence level. The results suggest that our model is robust and is applicable for COVID–19 vaccine hesitancy models in other countries.

## Data Availability

The data can be accessed via https://www.worldometers.info/coronavirus/country/us/

## Notes

### Competing Interest Statement

The authors have declared no competing interest.

